# A formula for the basic reproduction number of an infectious disease in a heterogeneous population with structured mixing

**DOI:** 10.64898/2026.03.27.26349419

**Authors:** Ewan Colman, Anastasia Chatzilena, Bastian Prasse, Leon Danon, Ellen Brooks-Pollock

## Abstract

The basic reproduction number of an infectious disease is known to depend on the structure of contacts between individuals in a population. This relationship has been explored mathematically through two well-known models: one which depends on a matrix of contact rates between different demographic groups, and another which depends on the variability of contact rates over the population. Here we introduce a model that combines and generalises these two approaches. We derive a formula for the basic reproduction number and validate it through comparisons to simulated outbreaks. Applying this method to contact survey data collected in Belgium between 2020 and 2022, we find that our model produces higher estimates of the basic reproduction number and larger relative changes over periods when social contact behaviour was changing during the COVID-19 pandemic. Our analysis suggests some practical considerations when using contact data in models of infectious disease transmission.

---

Many studies in mathematical epidemiology explore how the structure of contacts within a population effects the spread of disease [1–3]. In this context, *contact* can be any interaction capable of causing the disease to transmit. With data about contact behaviour becoming increasingly available, much of the theory is proving to be useful in the real world [4–9]. This was particularly true during the height of the COVID-19 pandemic when policy makers relied heavily on models to predict the consequences of imposing restrictions on the population [10, 11].

The relationship between contact and the spread of infectious disease is often understood through the *basic reproduction number, R*_0_, defined as the number of secondary cases caused by a typical infected individual in an otherwise susceptible population [12, 13]. Efforts to control a disease often focus on ensuring that *R*_0_ *<* 1, as this threshold determines when major outbreaks are possible. There has therefore been considerable attention given to how *R*_0_ is calculated, and in particular, the meaning of the word “typical” in the definition above.

Naively, *R*_0_ can be calculated by multiplying the mean number of contacts an individual has during their infectious period by the probability that contact results in transmission. This, however, assumes that each individual in the population is equally likely to be infected, when in reality patterns of connectivity can cause some individuals to be more exposed than others and therefore more likely to be transmitters of the disease. More specifically, the simple calculation assumes that the population is both *well-mixed*, meaning anyone can potentially engage in contact with anyone else, and *homogeneous* with regard to contact rates, meaning all individuals behave the same [14]. There are two notable ways in which the calculation has been adapted to relax these assumptions.

The approach introduced by Nold, and later Diekmann et al, considers a population stratified into distinct groups (e.g. age bands) and constructs a *next-generation matrix* that estimates the infection pressure that each group exerts on each other group [15, 16]. An alternative method introduced by Anderson et al considers variability between individuals in terms of the number of contacts they have [17]. We refer these model assumptions as *group-dependent mixing* and *heterogeneous contact rates*, respectively. Fig. 1 illustrates how these models capture two distinctly different features of contact behaviour.

**Figure 1:**
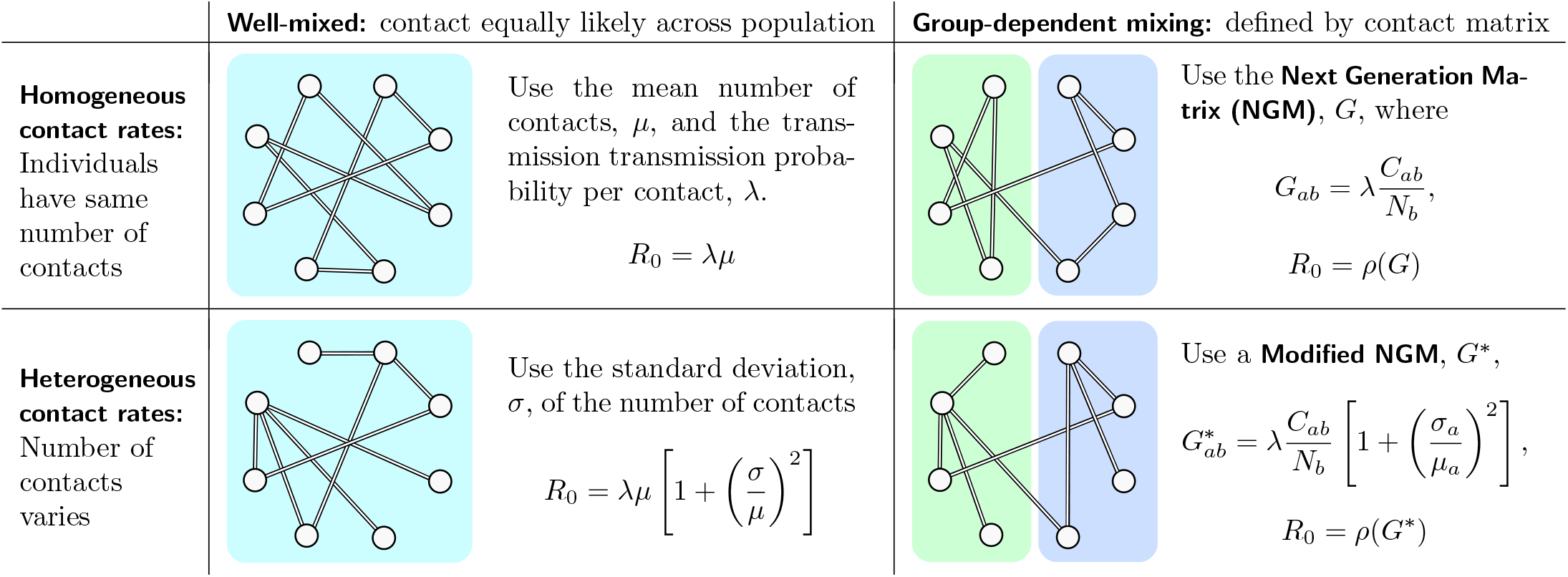
Recipe for calculating the basic reproduction number depending on contact patterns in the underlying population. Model shown in the top row and left column are known results. The result on the bottom right is derived in Section 1 and shown to generalise the other models. Indices *a* and *b* refer to the groups into which the population are partitioned, *C*_*ab*_ is the number of contacts between groups *a* and *b, N*_*b*_ is the population of group *b, µ*_*a*_ and *σ*_*a*_ are the mean and standard deviation for the set of individuals in *a, ρ*(·) is the spectral radius (largest eigenvalue) of the matrix

Both methods have found applications in the real world. For example, questionnaire surveys asking participants to give information about their daily contacts provide sufficient data for *R*_0_ to be calculated using either approach [18, 19]. Furthermore, both methods were utilised during the COVID-19 pandemic and used as effective instruments to effectively communicate the potential impact of non-pharmaceutical interventions to policy makers [20–23]. A concern here is that different conclusions may be reached, even when the underlying data are the same.

In this paper, we introduce a model that incorporates *both* the mixing patterns between different groups in the population *and* the heterogeneity in individual rates of contact (lower right panel of Fig. 1). We derive the basic reproduction number and show that our model is a generalisation of the two established methods. Our results are validated against simulated outbreaks and we provide a demonstration of how such methods should be applied using contact survey data collected in Belgium during the pandemic.

## 1 Deriving *R*_0_ in a population with group-dependent mixing and heterogeneous contact rates

In this section we derive the novel formula for *R*_0_ shown in the lower right hand quadrant of Fig. 1. We then show that this formula generalises other common approaches to calculating *R*_0_ shown in the three other quadrants of Fig. 1.

### 1.1 Network construction

To model the effects of both group-dependent mixing and contact heterogeneity, we consider a population of *N* individuls stratified into *m* groups, indexed by *a* = 1, 2, …, *m*, and that individuals can be identified by their number of contacts *k* = 1, 2, …, *K* where *K* is an arbitrary upper limit. We use *c*_*ab*_ to denote the number of contacts between people in group *a* and group *b*, and *n*_*k,a*_ to denote the number of individuals in group *a* with exactly *k* contacts (for all *a* = 1, 2, …, *m, b* = 1, 2, …, *m* and *k* = 1, 2, …, *K*). We use *c*_*a*_ to denote the total number of contacts of those in group *a* (for all *a* = 1, 2, …, *m*), thus

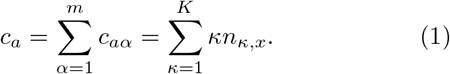

The underlying individual-based network may be modelled as the expected value of a degree-corrected stochastic block model [24], corresponding to an *N* × *N* adjacency matrix *A* whose entries equal the probability of a link between the respective nodes. The probability of a contact existing from node *i* to node *j* is constructed by taking the product of three things: the number of contacts of node *i*, the fraction of *i*’s contacts that are in the group that contains *j*, and the fraction of contacts in the group of *j* that connect to node *j*. Supposing *i* is in group *a* with *k* contacts and *j* is in group *b* with *l* contacts, we obtain the probability of a link between node *i* and *j* as

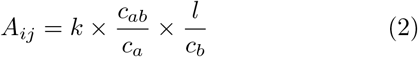

Note that *A*_*ij*_ = *A*_*ji*_.

The partition of the network into *m* × *K* cells with respect to group of each node and its number of contacts is *equitable*, meaning that all nodes within the same cell are identical in the way they share contacts with other cells [25]. Letting *C*_*p*_ denote the set of nodes in cell *p* where *p* = 1, 2, …, *K* correspond to the cells containing individuals in group 1 with *k* = 1, …, *K* contacts, respectively, *p* = *K* +1, …, 2*K* correspond to group 2 with *k* = 1, 2, …, *K* contacts, and so on (in general, the index *p* corresponds to individuals in group *a* = ⌈*p/K* ⌉ with *k* = 1 + (*p* − 1 mod *K*) contacts), we have that *A* = *XQX*^*T*^ where *X* is a matrix of size *N* × *mK* with orthonormal columns and

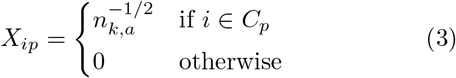

and *Q* is an is a matrix of size *mK* × *mK*, known as the *quotient matrix* of the equitable partition, whose elements are

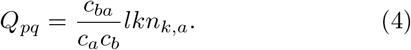

Suppose we have an infectious disease spreading over the contacts of the network. We define *λ* to be the probability that an infected individual, at some point during their period of being infected, infects a given susceptible neighbour (an adjacent node). It is known that if the infection probability satisfies *λ <* 1*/ρ*(*A*), where *ρ*(*A*) is the dominant eigenvalue of the adjacency matrix *A*, a major outbreak does not occur [26,27]. Furthermore, if the network has an equitable partition, *ρ*(*A*) = *ρ*(*Q*) [25].

### 1.2 Next generation matrix

Let *G*_*pq*_ be the expected number of infections in *C*_*p*_ caused by each infected individual in *C*_*q*_. The matrix *G* is known as the *next generation matrix* (NGM). Since transmission occurs on each contact with probability *λ*, we have

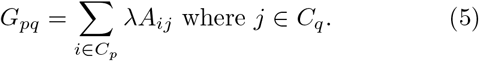

Substituting Eq.(2) into the equation above and simplifying, we see that *G* = *λQ*. It is known that the largest eigenvalue of *G* is equal to *R*_0_ [15,16]. Since *ρ*(*G*) = *λρ*(*Q*), the threshold criteria that major outbreaks cannot occur if *R*_0_ *<* 1 is exactly equivalent to the threshold criteria mentioned above.

### 1.3 Eigenvalues of the NGM

To save space, we define *M*_*ab*_ = *c*_*ab*_*/*(*c*_*a*_*c*_*b*_) for all the *a*’s and *b*’s, so *G*_*pq*_ = *λlM*_*ba*_ × *kn*_*k,a*_. The NGM can be written as the product of matrices of dimension *mK* × *m* and *m* × *mK*

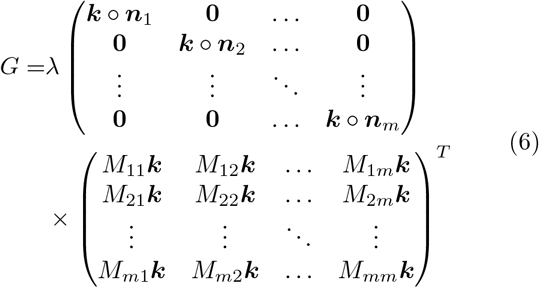

where

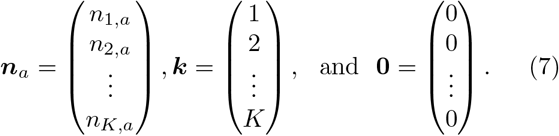

The ◦ symbol has been used to denote element-wise multiplication.

We make use of the fact that the eigenvalues of the product of two matrices are unchanged if the order of multiplication is reversed (***v*** = *B****u*** transforms *e****v*** = *AB****v*** into *e****u*** = *BA****u***) Reversing the product in (6) produces an *m* × *m* matrix whose *a, b*th entry is

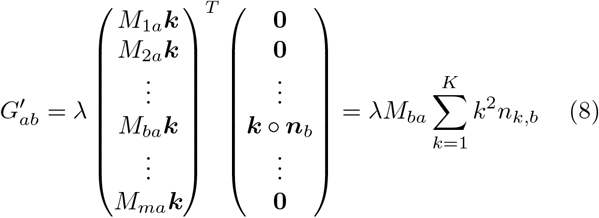

Introducing *µ*_*a*_ and 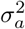 to denote the mean and variance of the number of contacts for individuals in group *a* for all *a* = 1, 2, …, *m*, we can rewrite the above as

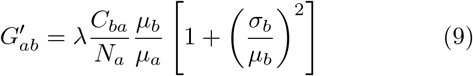

where *N*_*a*_ denotes the number of individuals in group *a*. We simplify further by noting that the matrix *G*^′^ has exactly the same eigenvalues as the matrix *G*^∗^ = [*DG*^′^*D*^−1^]^*T*^ where *D* is a diagonal matrix with entries *µ*_1_, *µ*_2_, …, *µ*_*m*_. The *a, b*th entry is

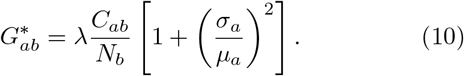

Since *G*^∗^ has precisely the same eigenvalues as the NGM, *R*_0_ = *ρ*(*G*^∗^). Noting the similarity between this matrix and the definition of the next-generation matrix in the case of *homogeneous contact rates*, we refer to *G*^∗^ as the *modified next generation matrix*.

### 1.4 Special cases

The result from Diekmann et al [16], corresponding to a model with *group-dependent mixing* and *homogeneous contact rates*, is recovered when we assume no variance in the number of contacts within each group. Setting *σ*_*a*_ = 0 for all *a*,

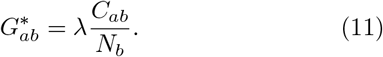

The result originally from Anderson et al [17], and derived elsewhere through other methods [23, 28, 29], corresponding to a model that is *well-mixed* with *heterogeneous contact rates*, is recovered by assuming that contact happens independently of the group mixing structure, hence

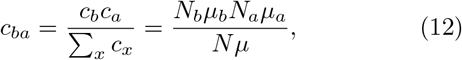

where *N* and *µ* are the total population size and the population average number of contacts, respectively. Substituting this into Eq.(10) gives

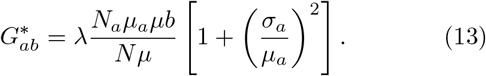

We can then write *G*^∗^ as an outer product

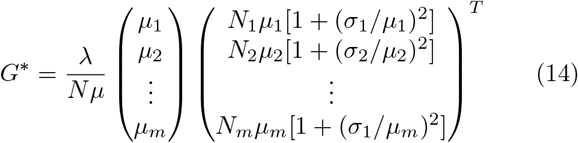

Following [30], the largest eigenvalue, *R*_0_, is found by calculating the inner product

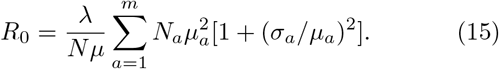

From the definition of variance, 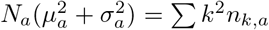, which transforms the summation above to ∑*k*^2^ *n*_*k*_ = *N* (*µ*^2^ + *σ*^2^), where *n*_*k*_ = ∑*n*_*k,a*_, giving

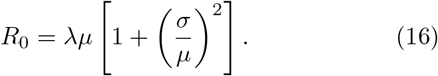

Finally, when setting *σ* = 0 we arrive at the result which corresponds to a population that is *well-mixed* with *homogeneous contact rates, R*_0_ = *λµ*, as used in early models [14, 31].

### 1.5 Validation against simulated outbreak

We validate the analytically derived *R*_0_ by comparing it to simulated outbreaks. We start by constructing synthetic contact networks on which we will simulate outbreaks of an infectious agent. Two parameters determine the topology of the network: *h* is the contact heterogeneity, and *m* is the level to which one group dominates contact. We implement a particular procedure for constructing networks that allows these two parameters to be varied while holding other structural properties approximately equal. This analysis does not validate the model on all possible networks, but does allow for examination of our results over a range of network topologies.

Nodes are assigned to two groups, *A* and *B*, of equal size. Let *E* be the number of edges. In our construction, the number of edges between *A* and *A* is equal to the number between *A* and *B*, and is the nearest whole number to *E*(1 − *m*)*/*3. The remaining edges are between *B* and *B*. When *m* = 0, there is no dependence on group membership. As *m* → 1 an increasing proportion of the edges are between *B* and *B*. This dependence is illustrated in the three figures parallel to the horizontal axis in Figure 2.

**Figure 2:**
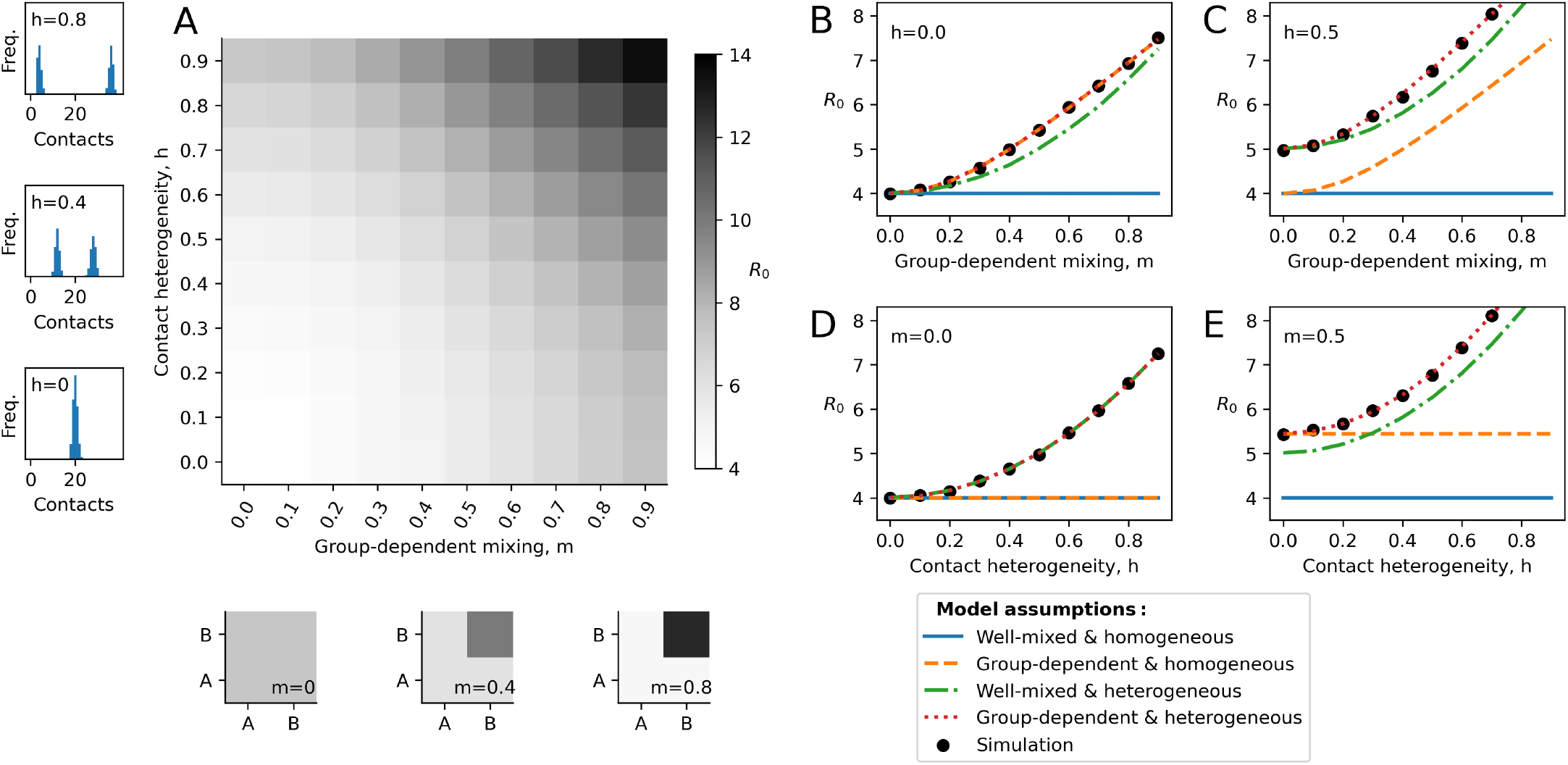
Comparison of the basic reproduction number in a simulated epidemic data and four methods for its calculation. A. The reproduction number obtained from simulated outbreaks on a network parameterized by heterogeneity in the number of contacts and strength of group-dependent mixing. Smaller figures aligned with the axes illustrate how parameter values affect the contact structure. B. Dependence of the basic reproduction number on the level of group-dependent mixing and the method of calculation. In this case contact heterogeneity is not present in the network other than the difference between group B and group A (when *m >* 0). C. Equivalent to panel B but heterogeneity between contacts within and between groups. D. Dependence on contact heterogeneity for networks with no explicit group mixing. E. Similar to D but with group-dependent mixing. The combined model aligns with simulation outcomes in all parameterizations of the contact network.

Each group is further divided equally into high degree and low degree classes. The number of edges between high degree nodes in group *X* and high degree nodes in *Y*, where *X* and *Y* can be either *A* or *B*, is *E*_*XY*_ (1 + *h*)^2^*/*4. Between high degree nodes in group *X* and low degree nodes in *Y* it is *E*_*XY*_ (1 + *h*)(1 − *h*)*/*4 (note if *X* = *Y* then this quantity is doubled), and between low degree nodes in group *X* and low degree nodes in *Y* is *E*_*XY*_ (1 − *h*)^2^*/*4. Partitioning the edges this way leaves the total number of edges between each pair of groups fixed. When *h* = 0, all nodes have the same degree, as *h* → 1 the degree of the low degree class approaches 0 and all the connectivity is between the high degree nodes. The dependence of the degree distribution on *h* is illustrated in the three figures parallel to the vertical axis in Figure 2.

We simulate an outbreak of an infectious agent on the network. To be consistent with the assumptions of the NGM model, nodes do not acquire immunity after becoming infected. One run of the simulation goes as follows: let ℐ_*g*_ be the nodes infected in generation *g*. To begin, ℐ_*g*_ is a set containing one randomly selected node. For a fixed number of generations, ℐ_*g*_ is constructed by selecting each neighbour of each nodes in ℐ_*g*−1_ with probability *λ*. The computed *R*_0_ is |ℐ_*g*_ |*/*|ℐ_*g*−1_| for some value of *g* large enough for the value at each generation to have converged.

Figure 2 panel A shows the outcome of these simulations over a range of network topologies. All simulations were performed on networks with 10^4^ nodes, 10^5^ edges, and *λ* = 0.2. We present the value of *R*_0_ computed at *g* = 7 (if the outbreak had ended before the 7th generation then it is discarded). We see that heterogeneity, in terms of both the distribution of the numbers of contacts (*h*) and the mixing matrix (*m*) contribute to larger values of *R*_0_.

Panels B-E compare simulation outcomes to analytical results. In all cases, the method built on a group-dependent and heterogeneous model, introduced in section 1.3, shows good agreement with simulated outcomes. The method built on a group-dependent and homogeneous model performs well when degree heterogeneity is not present in the simulation (*h* = 0). Similarly, the model that does not explicitly include the effects of group-mixing performs well when group-mixing is not present in the simulation (*m* = 0).

## 2 Application to contact survey data

### 2.1 Data

We use the CoMix contact survey in Belgium collected between April 2020 and March 2022 [10]. Surveys were completed over 43 waves approximately 2 weeks apart. Each wave had a recruitment target of 1500 participants with an aim to retain as many participants from previous waves as possible. Among the questions, participants were asked to give their own age and estimate the ages of all individuals they had contact with on the day prior to completing the survey.

Data imputation needed to be performed to be able to group participants and their contacts in into distinct age groups. The ages of participants were given in 10 distinct age groups. The estimated ages of contacts were given as ranges of possible values. For each one, an exact age was assigned to it by choosing randomly from the range of possible values with probability weighted by the population age distribution (according to World Population Prospect estimates for 2020 [32]). Contacts were then assigned to the 10 age groups according to these imputed ages.

Each wave of the study is analysed separately. Bootstrapping is used to quantify the uncertainty that arises as a result of variability in the process recruiting participants randomly from the population. For each one, 10^4^ bootstrap samples are created by sampling with replacement from the set of study participants until the total number sampled is equal in size to the original. In such a sample, a participant selected multiple times is considered to represent multiple different individuals from the population.

The quantities *N*_*k,a*_ (for all numbers of contacts *k*, and age groups *a*) are computed by first counting survey participants in age group *a* that have exactly *k* contacts, and then multiplying by the appropriate factor to correct for uneven sampling across the different age groups: *Q*_*a*_ = *N*_*a*_*/P*_*a*_ where *N*_*a*_ is the total population of people in age group *a* and *P*_*a*_ is the number or participants in age group *a* in the wave.

The quantities *c*_*a,b*_ (for all combinations of age groups *a* and *b*) are computed by first counting the total number of contacts between participants in group *a* and their reported contacts in group *b*, for all *a* and *b*. Suppose this raw count is denoted 𝒞_*ab*_. We use *c*_*ab*_ = (𝒞_*ab*_*Q*_*a*_ + 𝒞_*ba*_*Q*_*b*_)*/*2. This method is used to correct for uneven sampling across the population and also ensure that *c*_*ab*_ = *c*_*ba*_ [33].

### 2.2 Removal of outliers

The values of *R*_0_ calculated on these contact data can be dominated by a small number of individuals reporting unusually large numbers of contacts. For this reason we omit participants who report a number of contacts that is abnormally large compared to the majority. As contact distributions tend to be highly right-skewed [34], we choose to work with the log-transformed distribution of number of contacts. We calculate the mean and standard deviation for the natural logarithm of the number of contacts of all participants in the wave, and calculate the *z*-score (the number of standard deviations above the mean) for each participant. Those with a *z*-score above a given threshold, *z*, are omitted.

Panel A in figure 3 shows the outcome of calculating *R*_0_ with no outliers removed compared to the outcome when outliers are removed using a threshold of *z* = 3.5. Sensitivity to this threshold can be seen in Panel B. At *z* = 3.5, between 0 and 20 participants out of total of more than 1000 have been omitted. This small number of omissions has a substantial effect not just on the magnitude of *R*_0_, but also on the uncertainty obtained from bootstrapping and the relative changes seen over time. Three major events regarding Non-pharmaceutical interventions correspond to changes in contact patterns become visible when outlier removal is applied [35, 36].

**Figure 3:**
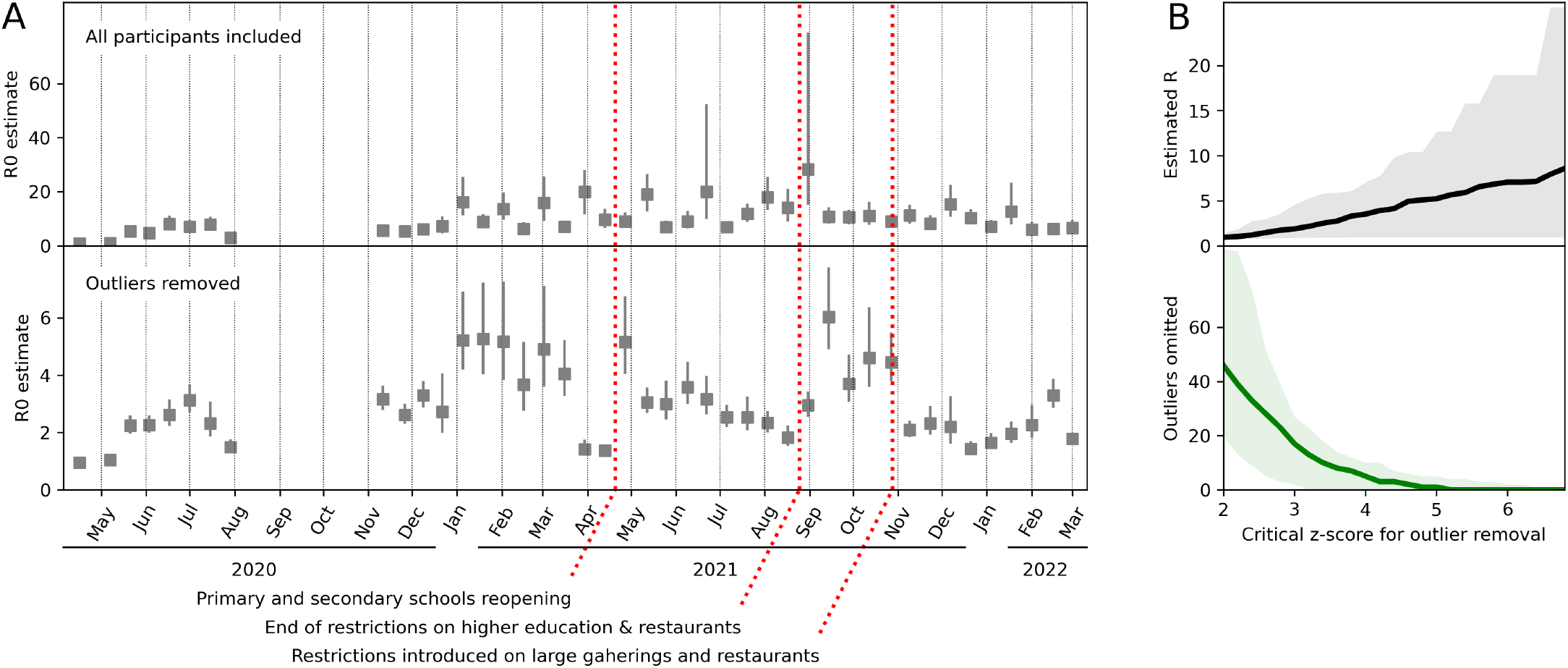
Estimated basic reproduction numbers and sensitivity to removal of outliers. A. The basic reproduction number calculated using the combined method (group-dependent & heterogeneous) for all waves of CoMix in Belgium. Median (squares) and 95% confidence intervals (lines) of 10^4^ bootstrap samples are shown. The lower panel shows outcomes after outliers have been removed using a threshold value of *z* = 3.5. Annotations show the time at which various changes in policy occurred. B. The effect of the threshold value on the basic reproduction number calculated using the combined method (upper panel) and the number of outliers removed from the data before making the calculation (lower panel). shaded areas show the range of outcomes over the 43 waves.

In general, a logical choice for *z* is a value where the expectation (according to some assumption about the underlying population) of the number of outliers omitted is less than 1. For example, if we were to assume that the distribution of contact numbers is log-normal, then from a sample of 1500, the expectation of the number of values greater than 3.5 is approximately 0.3. Therefore anyone removed is well outside the distribution.

### 2.3 Effect of changes in contact behavior

We compare two waves of the CoMix study in Belgium where the impact of non-pharmaceutical interventions creates an apparent difference in contact behaviour. Data from a period of high stringency (wave 20) are shown in Figure 4 panels A and B. Participants were invited to complete the questionnaire on April 13th 2021 and responses were accepted for 2 weeks. For the first 6 days of this period (when most of the responses were received) school classes in primary and secondary schools were suspended, for the first 12 days cafes and restaurants were closed. These non-pharmaceutical interventions had been lifted before the start of a low stringency period (wave 21), shown in panels C and D, which started on April 27th.

**Figure 4:**
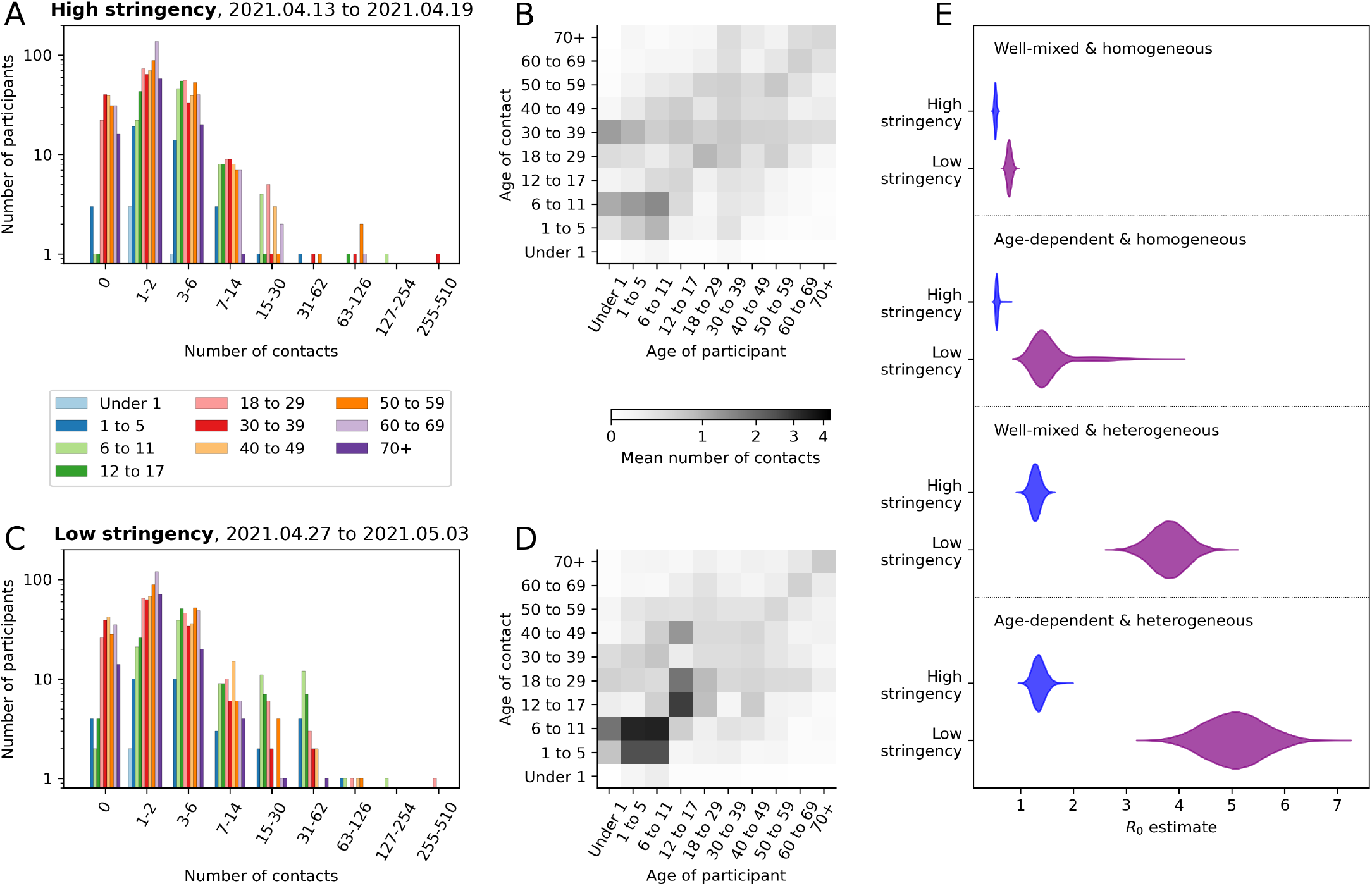
Comparison of methods applied to two waves of the CoMix study in Belgium. A. The number of participants reporting given number of contacts during a high stringency period (wave 20) when schools were closed, gyms and places of worship restricted. B. Age mixing matrix for the high stringency period (after adjusting to make consistent with population demographic data). C. Contact distribution during a low stringency period (wave 21) when schools were active and other restrictions relaxed. D. Age mixing matrix for the low stringency period. E. The distribution of reproduction numbers over 10^4^ bootstrap samples calculated using the four sets of model assumptions.

Consistent with the results from synthetic networks shown in Figure 2, methods that account for heterogeneity produce larger values of *R*_0_. In all four methods, the estimated *R*_0_ is larger for the low stringency period than the high stringency period, with some overlap in the bootstrap distributions for the two methods that assume homogeneous contact rates. When the well-mixed homogeneous model is applied, *R*_0_ increases by 50% from the high stringency period to the low stringency period, when the age-dependent homogeneous model is applied the increase is considerably higher, when the well-mixed heterogeneous model is applied the increase in *R*_0_ is higher still, and when the age-dependent model heterogeneous is applied *R*_0_ the increase is almost 300%. Thus, conclusions about the effect of NPIs may differ depending on the method used.

## 3 Discussion

The main contribution of this paper is a novel method for calculating the basic reproductive number of an infectious disease using data on the contact structure of the population. The approach builds on a well-known result in the field of mathematical epidemiology concerning the dependence of *R*_0_ on mixing rates between population subgroups, and another that links *R*_0_ to the amount of heterogeneity in individual contact rates across the population. The introduced method unifies and generalises these two established approaches. We have seen that the established methods fail to provide an accurate estimate of *R*_0_ compared to simulations when both group mixing and contact heterogeneity are present, whereas the introduced method successfully incorporates both features of population structure.

Our results carry considerable implications regarding the way contact survey data has been used to inform policy. Analysis of the CoMix study, that was regularly circulated within scientific advisory groups throughout the first few years of the COVID-19 pandemic, used the next generation matrix approach with an age-dependent stratification [10, 22, 37]. Other analyses chose to report the mean number of contacts to indicate the connection between the introduction or relaxation of NPIs and potential disease transmission [38]. Our findings challenge the validity of these methods as they fail to capture significant changes in transmission that can arise from changes to the distribution of contact rates.

For example, when bans on large gatherings are lifted, a minority of people increase their contacts while the majority remain approximately the same. In this situation, the mean number of contacts might only increase by a small amount whereas heterogeneity, and therefore *R*_0_, could increase substantially. Several models built for quick policy-focused analysis did recognise the importance of including heterogeneity [20, 21, 23]. However, neglecting the role of age-dependent mixing may invite valid criticism, particularly considering that school closures - one of the most controversial measures implemented - affected mixing in a very age-dependent way. Our results suggest that age-dependent mixing plays a relatively minor part in determining *R*_0_, but there is little reason to debate this now that we have provided a combined method. That said, there are some limitations of our approach which we now point out.

While our method accurately predicted the outcome of simulated outbreaks, the simulation was designed to follow the assumptions of the next generation matrix model and is therefore not an entirely accurate representation of reality. Notably, there is no concept of immunity in the model, which is known to have an effect even during the early stages of an outbreak [39, 40]. Additionally, we chosen a range of networks that expose the importance of group mixing and contact heterogeneity, however, other features of contact behaviour may also be relevant to the spread of disease. This has recently been shown with respect to degree assortativity in networks using a model with two levels of social connectivity [41].

Another limitation is our assumption that all individuals in the same group with the same number of contacts are distributing their contacts in a similar way. To give an example, suppose Alice and Bob are both in group *A*, Alice has *k* contacts in group *A* and 0 contacts in *B*, Bob has *k* contacts in *B* and 0 in *A*, in our model this would be indistinguishable from a situation where Alice and Bob both have *k/*2 contacts in *A* and *k/*2 in *B*. If this situation were to occur in real world data it would perhaps suggest the need to use additional variables such as profession to stratify the population.

In addition to the mathematical contribution, we have provided some analysis of real-world contact data to demonstrate how these methods can be applied. An important step in this process is the removal of data points that may be considered outliers. In doing so we are implicitly saying that contact data are in some way incorrect, possibly suggesting that some contact survey participants have not interpreted the question in the way the questionnaire designers intended. A future direction for this research would be to incorporate a deeper understanding of how social networks form and evolve over time [42–44].

We conclude by making some simple recommendations. Firstly, the method developed in this paper should be adopted by anyone using contact surveys to obtain estimates of *R*_0_. As we saw during the COVID-19 pandemic, these methods provide instant modelling outputs that can be used to engage policy makers with the modelling process; the unified method will avoid arriving at contradictory results. Secondly, work needs to be done to obtain high accuracy information about individuals with very large numbers of contacts. Their role in superspreading is long known to be a major contributor to disease dynamics [45]. Lastly, this area of research can be developed further by incorporating a wider range of data sources, temporal variables, and cross-disciplinary research into social behaviour.

## Data Availability

All data produced are available online at https://github.com/EwanColman/Reproduction-number-in-
heterogeneous-populations-with-structured-mixing

https://github.com/EwanColman/Reproduction-number-in-heterogeneous-populations-with-structured-mixing

## Acknowledgments

We are grateful to all involved with the collection and curation of the CoMix data, especially Pietro Coletti for quickly responding to our inquiries and requests. We thank Disa Hansson of ECDC for helpful feedback on early results from this work. For the purpose of open access, the author(s) has applied a Creative Commons Attribution (CC BY) licence to any Author Accepted Manuscript version arising from this submission.

This project was supported by a Research England grant awarded to University of Bristol. EBP acknowledges the support of the National Institute for Health Research Health Protection Research Unit (NIHR HPRU) in Evaluations and Behavioural Science at the University of Bristol. LD is funded by UK Research and Innovation AI programme of the Engineering and Physical Sciences Research Council (EPSRC grant EP/Y028392/1). LD and EBP are affiliated with the JUNIPER partnership funded by the Medical Research Council (MRC grant MR/X018598/1; https://www.ukri.org/councils/mrc/) and acknowledge the support of the National Institute for Health Research Health Protection Research Focus Award in Immunisation at the University of Bristol.

## Data access statement

No new data were created during this study.

## References

[1] Matt J Keeling and Pejman Rohani. Modeling infectious diseases in humans and animals. Princeton university press, 2011.

[2] István Z Kiss, Joel C Miller, Péter L Simon, et al. Mathematics of epidemics on networks. Cham: Springer, 598(2017):31, 2017.

[3] Romualdo Pastor-Satorras, Claudio Castellano, Piet Van Mieghem, and Alessandro Vespignani. Epidemic processes in complex net-works. Reviews of modern physics, 87(3):925, 2015.

[4] Jacco Wallinga, Peter Teunis, and Mirjam Kretzschmar. Using Data on Social Contacts to Estimate Age-specific Transmission Parameters for Respiratory-spread Infectious Agents. American Journal of Epidemiology, 164(10):936–944, 09 2006.

[5] Kiesha Prem, Alex R. Cook, and Mark Jit. Projecting social contact matrices in 152 countries using contact surveys and demographic data. PLOS Computational Biology, 13(9):1–21, 09 2017.

[6] Andria Mousa, Peter Winskill, Oliver John Watson, Oliver Ratmann, Mélodie Monod, Marco Ajelli, Aldiouma Diallo, Peter J Dodd, Carlos G Grijalva, Moses Chapa Kiti, Anand Krishnan, Rakesh Kumar, Supriya Kumar, Kin O Kwok, Claudio F Lanata, Olivier Le Polain de Waroux, Kathy Leung, Wiriya Mahikul, Alessia Melegaro, Carl D Morrow, Jöel Mossong, Eleanor FG Neal, D James Nokes, Wirichada Pan-ngum, Gail E Potter, Fiona M Russell, Siddhartha Saha, Jonathan D Sugimoto, Wan In Wei, Robin R Wood, Joseph Wu, Juanjuan Zhang, Patrick Walker, and Charles Whittaker. Social contact patterns and implications for infectious disease transmission – a systematic review and meta-analysis of contact surveys. eLife, 10:e70294, nov 2021.

[7] Thang Hoang, Pietro Coletti, Alessia Melegaro, Jacco Wallinga, Carlos G Grijalva, John W Edmunds, Philippe Beutels, and Niel Hens. A systematic review of social contact surveys to inform transmission models of close-contact infections. Epidemiology, 30(5):723–736, 2019.

[8] D Mistry, M Litvinova, A Pastorey Piontti, M Chinazzi, L Fumanelli, MFC Gomes, SA Haque, QH Liu, K Mu, X Xiong, et al. Inferring high-resolution human mixing patterns for disease modeling. arxiv. arXiv preprint 2003.01214, 2020.

[9] JM Read, WJ Edmunds, S Riley, J Lessler, and DAT Cummings. Close encounters of the infectious kind: methods to measure social mixing behaviour. Epidemiology & infection, 140(12):2117–2130, 2012.

[10] Amy Gimma, James D. Munday, Kerry L. M. Wong, Pietro Coletti, Kevin van Zandvoort, Kiesha Prem, CMMID COVID-19 working group, Petra Klepac, G. James Rubin, Sebastian Funk, W. John Edmunds, and Christopher I. Jarvis. Changes in social contacts in england during the covid-19 pandemic between march 2020 and march 2021 as measured by the comix survey: A repeated crosssectional study. PLOS Medicine, 19(3):1–25, 03 2022.

[11] Kiesha Prem, Yang Liu, Timothy W Russell, Adam J Kucharski, Rosalind M Eggo, Nicholas Davies, Stefan Flasche, Samuel Clifford, Carl AB Pearson, James D Munday, et al. The effect of control strategies to reduce social mixing on outcomes of the covid-19 epidemic in wuhan, china: a modelling study. The lancet public health, 5(5):e261–e270, 2020.

[12] Johan Andre Peter Heesterbeek. A brief history of r 0 and a recipe for its calculation. Acta biotheoretica, 50(3):189–204, 2002.

[13] Matt J. Keeling and Bryan T. Grenfell. Individual-based perspectives on r0. Journal of Theoretical Biology, 203(1):51–61, 2000.

[14] James A. Yorke, Herbert W. Hethcote, and Annett Nold. Dynamics and control of the transmission of gonorrhea. Sexually Transmitted Diseases, 5(2):51–56, 1978.

[15] Annett Nold. Heterogeneity in disease-transmission modeling. Mathematical Biosciences, 52(3):227–240, 1980.

[16] Odo Diekmann, Johan Andre Peter Heesterbeek, and Johan Anton Jacob Metz. On the definition and the computation of the basic reproduction ratio r 0 in models for infectious diseases in heterogeneous populations. Journal of mathematical biology, 28:365–382, 1990.

[17] R. M. Anderson, G. F. Medley, R. M. May, and A. M. Johnson. A Preliminary Study of the Transmission Dynamics of the Human Immunodeficiency Virus (HIV), the Causative Agent of AIDS. Mathematical Medicine and Biology: A Journal of the IMA, 3(4):229–263, 10 1986.

[18] Jöel Mossong, Niel Hens, Mark Jit, Philippe Beutels, Kari Auranen, Rafael Mikolajczyk, Marco Massari, Stefania Salmaso, Gianpaolo Scalia Tomba, Jacco Wallinga, Janneke Heijne, Malgorzata Sadkowska-Todys, Magdalena Rosinska, and W. John Edmunds. Social contacts and mixing patterns relevant to the spread of infectious diseases. PLOS Medicine, 5(3):1–1, 03 2008.

[19] Leon Danon, Jonathan M Read, Thomas A House, Matthew C Vernon, and Matt J Keeling. Social encounter networks: characterizing great britain. Proceedings of the Royal Society B: Biological Sciences, 280(1765):20131037, 2013.

[20] Ellen Brooks-Pollock, Jonathan M Read, Thomas House, Graham F Medley, Matt J Keeling, and Leon Danon. The population attributable fraction of cases due to gatherings and groups with relevance to covid-19 mitigation strategies. Philosophical Transactions of the Royal Society B, 376(1829):20200273, 2021.

[21] Ellen Brooks-Pollock, Jonathan M Read, Angela R McLean, Matt J Keeling, and Leon Danon. Mapping social distancing measures to the reproduction number for covid-19. Philosophical Transactions of the Royal Society B, 376(1829):20200276, 2021.

[22] Christopher I Jarvis, Kevin Van Zandvoort, Amy Gimma, Kiesha Prem, Petra Klepac, G James Rubin, and W John Edmunds. Quantifying the impact of physical distance measures on the transmission of covid-19 in the uk. BMC medicine, 18:1–10, 2020.

[23] Ellen Brooks-Pollock, Kate Northstone, Lorenzo Pellis, Francesca Scarabel, Amy Thomas, Emily Nixon, David A Matthews, Vicky Bowyer, Maria Paz Garcia, Claire J Steves, et al. Voluntary risk mitigation behaviour can reduce impact of sars-cov-2: a real-time modelling study of the january 2022 omicron wave in england. BMC medicine, 21(1):25, 2023.

[24] Brian Karrer and Mark EJ Newman. Stochastic blockmodels and community structure in networks. Physical Review E—Statistical, Nonlinear, and Soft Matter Physics, 83(1):016107, 2011.

[25] S. Bonaccorsi, S. Ottaviano, D. Mugnolo, and F. De Pellegrini. Epidemic outbreaks in networks with equitable or almost-equitable partitions. SIAM Journal on Applied Mathematics, 75(6):2421– 2443, 2015.

[26] Piet Van Mieghem, Jasmina Omic, and Robert Kooij. Virus spread in networks. IEEE/ACM Transactions On Networking, 17(1):1–14, 2008.

[27] Deepayan Chakrabarti, Yang Wang, Chenxi Wang, Jurij Leskovec, and Christos Faloutsos. Epidemic thresholds in real networks. ACM Trans. Inf. Syst. Secur., 10(4), January 2008.

[28] Romualdo Pastor-Satorras and Alessandro Vespignani. Epidemic spreading in scale-free networks. Phys. Rev. Lett., 86:3200–3203, Apr 2001.

[29] Lauren Ancel Meyers, Babak Pourbohloul, M.E.J. Newman, Danuta M. Skowronski, and Robert C. Brunham. Network theory and sars: predicting outbreak diversity. Journal of Theoretical Biology, 232(1):71–81, 2005.

[30] Julia R Gog, Edward M Hill, Leon Danon, and Robin N Thompson. Vaccine escape in a heterogeneous population: insights for sars-cov-2 from a simple model. Royal Society Open Science, 8(7):210530, 2021.

[31] Klaus Dietz. Transmission and control of arbovirus disease. In in Proc. SIMS Conf. on Epidemiology (eds. D. Ludwig and KL Cooke), page 104, 1975.

[32] United nations, department of economic and social affairs, population division (2022). world population prospects 2022, data sources. un desa/pop/2022/dc/no. 9.

[33] Petra Klepac, Adam J Kucharski, Andrew JK Conlan, Stephen Kissler, Maria L Tang, Hannah Fry, and Julia R Gog. Contacts in context: large-scale setting-specific social mixing matrices from the bbc pandemic project. MedRxiv, pages 2020–02, 2020.

[34] Shweta Bansal, Bryan T Grenfell, and Lauren Ancel Meyers. When individual behaviour matters: homogeneous and network models in epidemiology. Journal of the Royal Society Interface, 4(16):879– 891, 2007.

[35] European centre for disease prevention and control response measures database.

[36] Lorenzo Lionello, Debora Stranges, Tommi Karki, Emma Wiltshire, Chiara Proietti, Alessandro Annunziato, Josep Jansa, Ettore Severi, ECDC-JRC Response Measures Database working group, et al. Non-pharmaceutical interventions in response to the covid-19 pandemic in 30 european countries: the ecdc–jrc response measures database. Eurosurveillance, 27(41):2101190, 2022.

[37] Pietro Coletti, James Wambua, Amy Gimma, Lander Willem, Sarah Vercruysse, Bieke Vanhoutte, Christopher I Jarvis, Kevin Van Zandvoort, John Edmunds, Philippe Beutels, et al. Comix: comparing mixing patterns in the belgian population during and after lockdown. Scientific reports, 10(1):21885, 2020.

[38] Spi-m-o: Summary of further modelling of easing restrictions – roadmap step 3, 5 may 2021. https://www.gov.uk/government/publications/spi-m-o-summary-of-further-modelling-of-easing-restrictions-roadmap-step-3-5-may-2021/spi-m-o-summary-of-further-modelling-of-easing-restrictions-roadmap-step-3-5-may-2021. Accessed: 2024-09-25.

[39] Lorenzo Pellis, Frank Ball, and Pieter Trapman. Reproduction numbers for epidemic models with households and other social structures. i. definition and calculation of r0. Mathematical biosciences, 235(1):85–97, 2012.

[40] Joel C Miller. Percolation and epidemics in random clustered networks. Physical Review E, 80(2):020901, 2009.

[41] Tom Britton and Frank Ball. Improving the use of social contact studies in epidemic modeling. Epidemiology, 36(5):660–667, 2025.

[42] Ewan Colman, Vittoria Colizza, Ephraim M Hanks, David P Hughes, and Shweta Bansal. Social fluidity mobilizes contagion in human and animal populations. eLife, 10:e62177, jul 2021.

[43] Gerardo Iñiguez, Sara Heydari, János Kertész, and Jari Saramäki. Universal patterns in egocentric communication networks. Nature Communications, 14(1):5217, 2023.

[44] Pierre Deville, Chaoming Song, Nathan Eagle, Vincent D Blondel, Albert-László Barabási, and Dashun Wang.Scaling identity connects human mobility and social interactions. Proceedings of the National Academy of Sciences, 113(26):7047–7052, 2016.

[45] James O Lloyd-Smith, Sebastian J Schreiber, P Ekkehard Kopp, and Wayne M Getz. Superspreading and the effect of individual variation on disease emergence. Nature, 438(7066):355–359, 2005.

